# Digital markers of motor speech impairments in natural speech of patients with ALS-FTD spectrum disorders

**DOI:** 10.1101/2023.04.29.23289308

**Authors:** Sanjana Shellikeri, Sunghye Cho, Sharon Ash, Carmen Gonzalez-Recober, Corey T. McMillan, Lauren Elman, Colin Quinn, Defne A. Amado, Michael Baer, David J Irwin, Lauren Massimo, Chris Olm, Mark Liberman, Murray Grossman, Naomi Nevler

## Abstract

**Background and objectives:** Patients with ALS-FTD spectrum disorders (ALS-FTSD) have mixed motor and cognitive impairments and require valid and quantitative assessment tools to support diagnosis and tracking of bulbar motor disease. This study aimed to validate a novel automated digital speech tool that analyzes vowel acoustics from natural, connected speech as a marker for impaired articulation due to bulbar motor disease in ALS-FTSD.

**Methods:** We used an automatic algorithm called Forced Alignment Vowel Extraction (FAVE) to detect spoken vowels and extract vowel acoustics from 1 minute audio-recorded picture descriptions. Using automated acoustic analysis scripts, we derived two articulatory-acoustic measures: vowel space area (VSA, in Bark^2^) which represents tongue range-of-motion (size), and average second formant slope of vowel trajectories (F2 slope) which represents tongue movement speed. We compared vowel measures between ALS with and without clinically-evident bulbar motor disease (ALS+bulbar vs. ALS-bulbar), behavioral variant frontotemporal dementia (bvFTD) without a motor syndrome, and healthy controls (HC). We correlated impaired vowel measures with bulbar disease severity, estimated by clinical bulbar scores and perceived listener effort, and with MRI cortical thickness of the orobuccal part of the primary motor cortex innervating the tongue (oralPMC). We also tested correlations with respiratory capacity and cognitive impairment.

**Results:** Participants were 45 ALS+bulbar (30 males, mean age=61±11), 22 ALS-nonbulbar (11 males, age=62±10), 22 bvFTD (13 males, age=63±7), and 34 HC (14 males, age=69±8). ALS+bulbar had smaller VSA and shallower average F2 slopes than ALS-bulbar (VSA: |*d*|=0.86, *p*=0.0088; F2 slope: |*d*|=0.98, *p*=0.0054), bvFTD (VSA: |*d*|=0.67, *p*=0.043; F2 slope: |*d*|=1.4, *p*<0.001), and HC (VSA: |*d*|=0.73, *p*=0.024; F2 slope: |*d*|=1.0, *p*<0.001). Vowel measures declined with worsening bulbar clinical scores (VSA: R=0.33, *p*=0.033; F2 slope: R=0.25, *p*=0.048), and smaller VSA was associated with greater listener effort (R=-0.43, *p*=0.041). Shallower F2 slopes were related to cortical thinning in oralPMC (R=0.50, *p*=0.03). Neither vowel measure was associated with respiratory nor cognitive test scores.

**Conclusions:** Vowel measures extracted with automatic processing from natural speech are sensitive to bulbar motor disease in ALS-FTD and are robust to cognitive impairment.

## 1. INTRODUCTION

Amyotrophic lateral sclerosis – frontotemporal dementia spectrum disorders (ALS-FTSD) are a group of neurodegenerative diseases with mixed motor neuron and frontotemporal lobe degeneration that result in a range of symptoms, including progressive muscle weakness and atrophy, difficulty with speech and swallowing, and changes in behavior, cognition, and/or personality.^1^ The variable and overlapping symptoms can make diagnosis and management challenging, and highlight the need for specific and reliable assessment tools to improve patient outcomes. Bulbar motor disease, impacting speech and swallowing functions, is a hallmark symptom of ALS. While only 30% of ALS patients begin with bulbar symptoms at onset (bulbar-onset ALS), over 90% develop bulbar disease over disease course.^2^ The earliest and most frequent sequela of bulbar disease is dysarthria, a motor speech disorder from a loss of articulator muscle strength and control that could arise from upper and/or lower motor neuron degeneration. This typically results in poorly intelligible speech requiring increased listener effort to comprehend,^3^ which can lead to communication breakdown, social isolation, and deterioration in quality of life.^4^ Bulbar motor disease can also result in swallowing difficulties and greater aspiration risk, which are common causes of severe morbidity and mortality in this population.^5,6^ Timely diagnosis and monitoring of bulbar motor disease in ALS-FTSD is critical.

Across ALS clinics, patient-reported functional rating scales such as the bulbar subscale of the ALS Functional Rating Scale-revised (ALSFRS-r)^7^, are the most routinely used methods to assess bulbar motor disease. These questionnaires are subjective and capture bulbar impairment at a gross functional level; thus they are limited as markers for screening and tracking of disease progression. Speech is a real-world activity and complex motor skill; many studies suggest that it may be among the most informative modalities for assessing motor abnormalities.^8^ Emerging automatic digital speech analyses have shown promise in providing quantitative clinical measures of bulbar impairment in ALS using highly structured speech tasks. ^9,10^ Yet, these structured tasks show poor face validity with natural real-life speech and thus can be less sensitive to mild bulbar motor impairment in early disease and subtle changes over time.^11–13^ Furthermore, structured tasks can be difficult to collect in patients with comorbid cognitive impairments, which are up to 50% of patients with ALS,^1^ due to issues with task comprehension or impaired reading skills. We expect that automatic analysis of speech can be adapted to natural speech tasks, such as picture descriptions, and provide sensitive and informative measures for bulbar disease.

Though all speech production subsystems may be affected,^14^ the articulatory subsystem which is controlled mostly by the tongue, is especially vulnerable to motor neuron disease.^15,16^ Available automatic speech platforms in ALS most commonly measure speaking rate in words per minute (WPM) but this measure can be confounded by respiratory or cognitive impairments.^17,18^ Three dimensional (3D) movement tracking studies link impaired articulation in ALS to slower and smaller tongue movements.^15^ Here we developed a novel digital speech analysis tool that assesses the articulatory subsystem by analyzing vowel acoustics that are directly related to tongue movements (speed and size) as a marker of bulbar motor disease in ALS. Speech features at the phoneme-level (i.e., at the level of the speech sound, such as vowels) may better reflect articulatory motor control processes than utterance-level features like speaking rate.^19^ This study assessed vowel measures of natural speech as potential markers of bulbar disease in ALS-FTSD. We hypothesized that vowel articulation measures of natural speech will be selectively impaired in ALS-FTSD patients with bulbar motor disease, and would be related to clinical bulbar motor signs and symptoms, speech intelligibility, and degeneration in oral regions of the primary motor cortex.

## 2. METHODS

### 2.1. Participants

Participants were selected from the University of Pennsylvania Integrated Neurodegenerative Disease Database (INDD^20^) as of December 31st, 2022, based on a clinical diagnosis of ALS with or without cognitive/behavioural impairments, i.e., ALS-FTSD^1^, or bvFTD,^21^ English as a primary language, and availability of at least one digitized picture description speech sample. The first available sample was used for analysis. bvFTD patients with evidence of a motor syndrome across lifespan clinical data were excluded. We also included speech data from historical healthy controls (HC) without a known cognitive or motor disorder.

For ALS, inclusion criteria included availability of clinical motor data within 3 months of speech sampling (average time interval between motor exam and speech sampling = 0.66 + 1.18 months), which was used to stratify patients into those with and without bulbar motor disease, herein referred to as ALS+bulbar and ALS-nonbulbar. Specifically, a score < 12 on the ALSFRS-r bulbar score^22^; and/or a score > 0 on the Penn Upper Motor Neuron scale (PUMNs) bulbar score^23^ indicated presence of bulbar disease. We also collected forced vital capacity in seated position (%FVC) in ALS patients, which is closely linked to respiratory capacity. Other clinical tests included the Edinburgh Cognitive Assessment Scale – North American version (ECAS-NA)^24^, available in n=61 ALS patients (85%) within 9 months of speech sampling (average time between ECAS and speech sampling= 0.85 + 2.22 months), and the Mini Mental State Exam (MMSE), available in n=42 ALS (58%) and all bvFTD and HC within 9 months of recording (average time between MMSE and speech sampling= 2.80 + 5.67 months).

The final cohort included 72 ALS patients (of which n=16 with cognitive impairments based on published cut-offs on ECAS-NA or MMSE); 25 bvFTD patients; and 32 HC. Seven ALS patients had bulbar onset with initial symptoms in speech or swallowing, while the remaining patients had limb onset (35 arm-onset, 27 leg-onset) or social/ behavioural onset (n=3). Irrespective of symptom onset site, n=49 ALS patients were classified as ALS+bulbar and n=23 as ALS-nonbulbar. We verified that ALS-nonbulbar patients did not present with lower motor neuron bulbar signs on their neurological exam. Demographic and clinical characteristics of the final groups are summarized in Table 1.

**Table 1.**
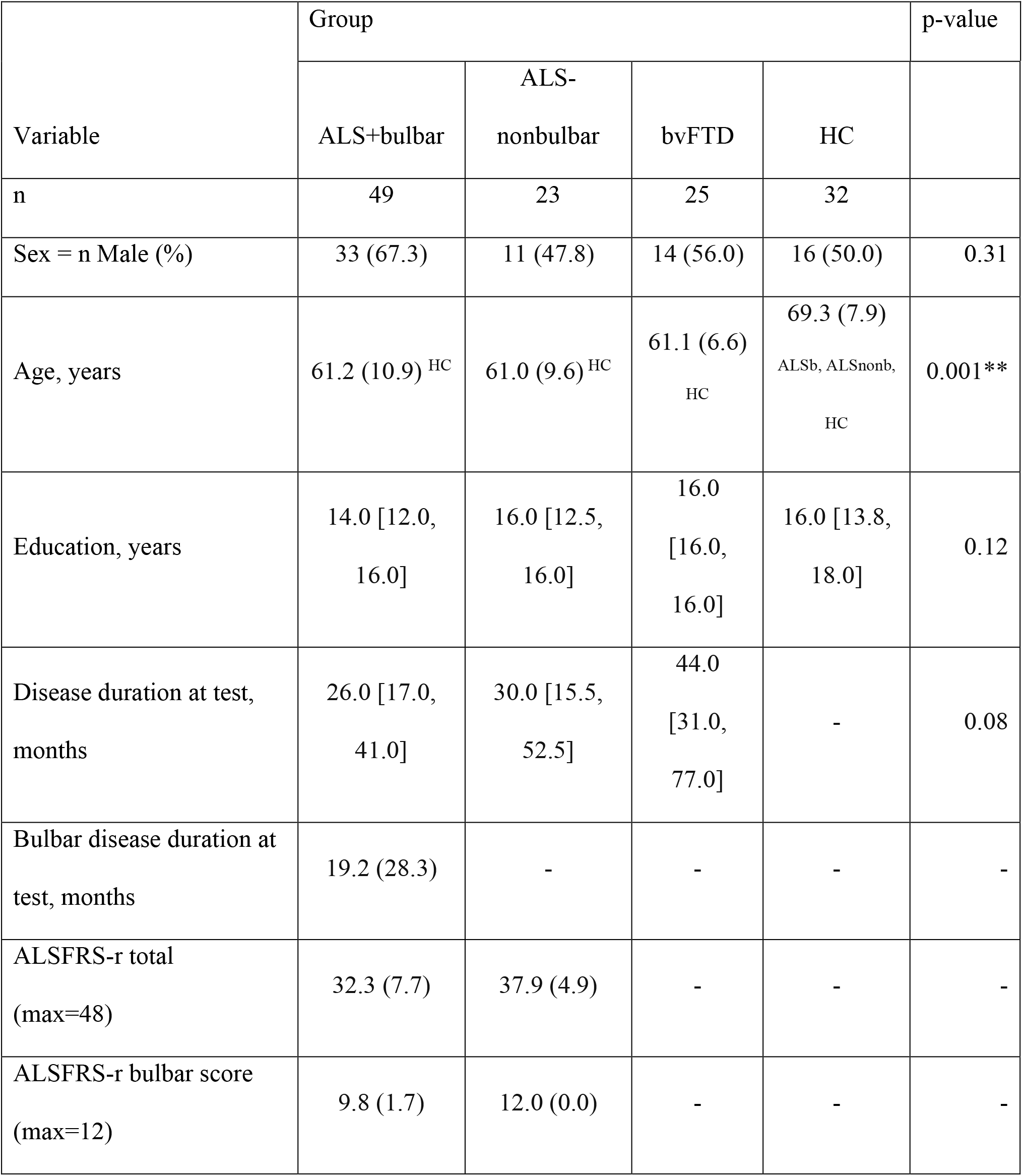
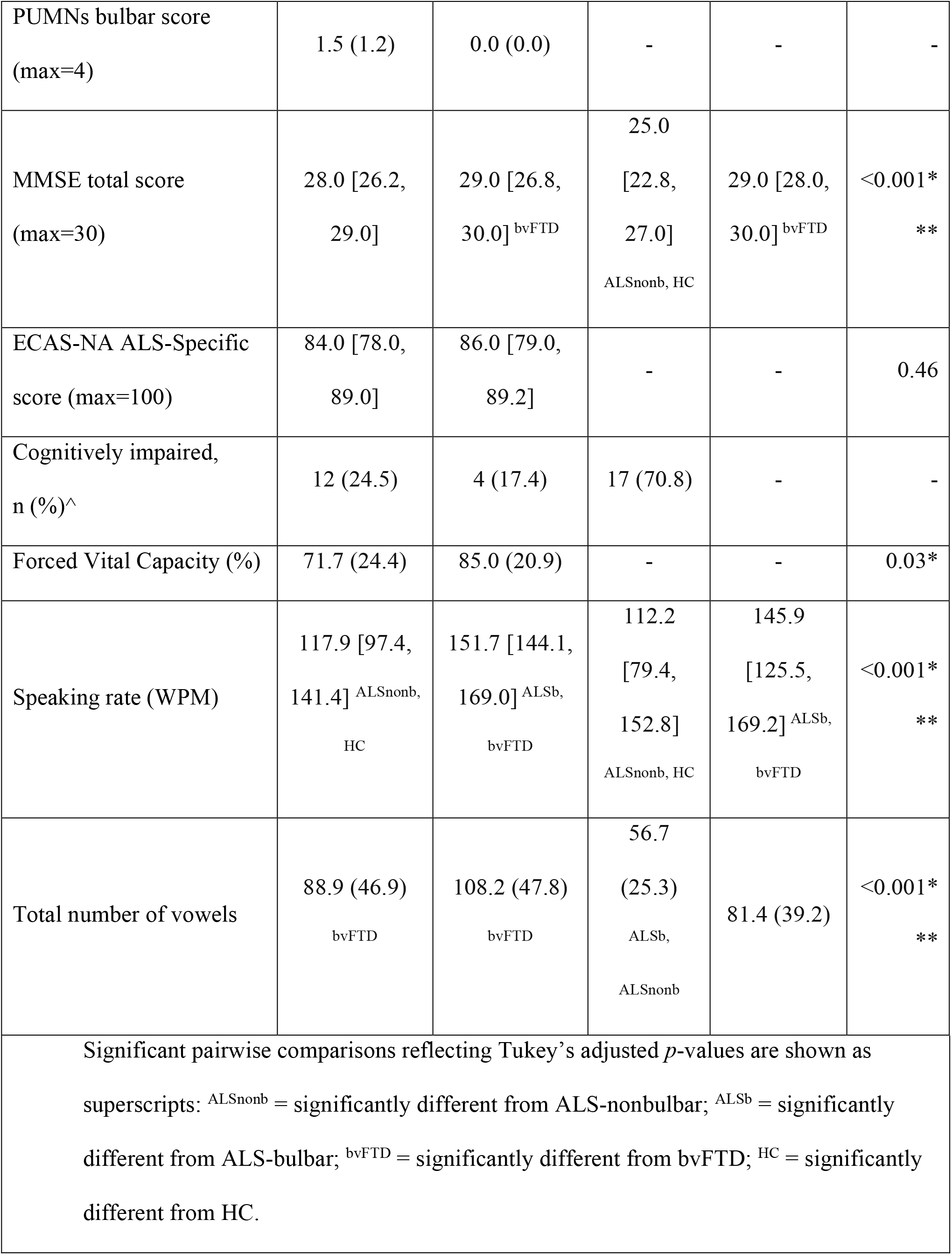
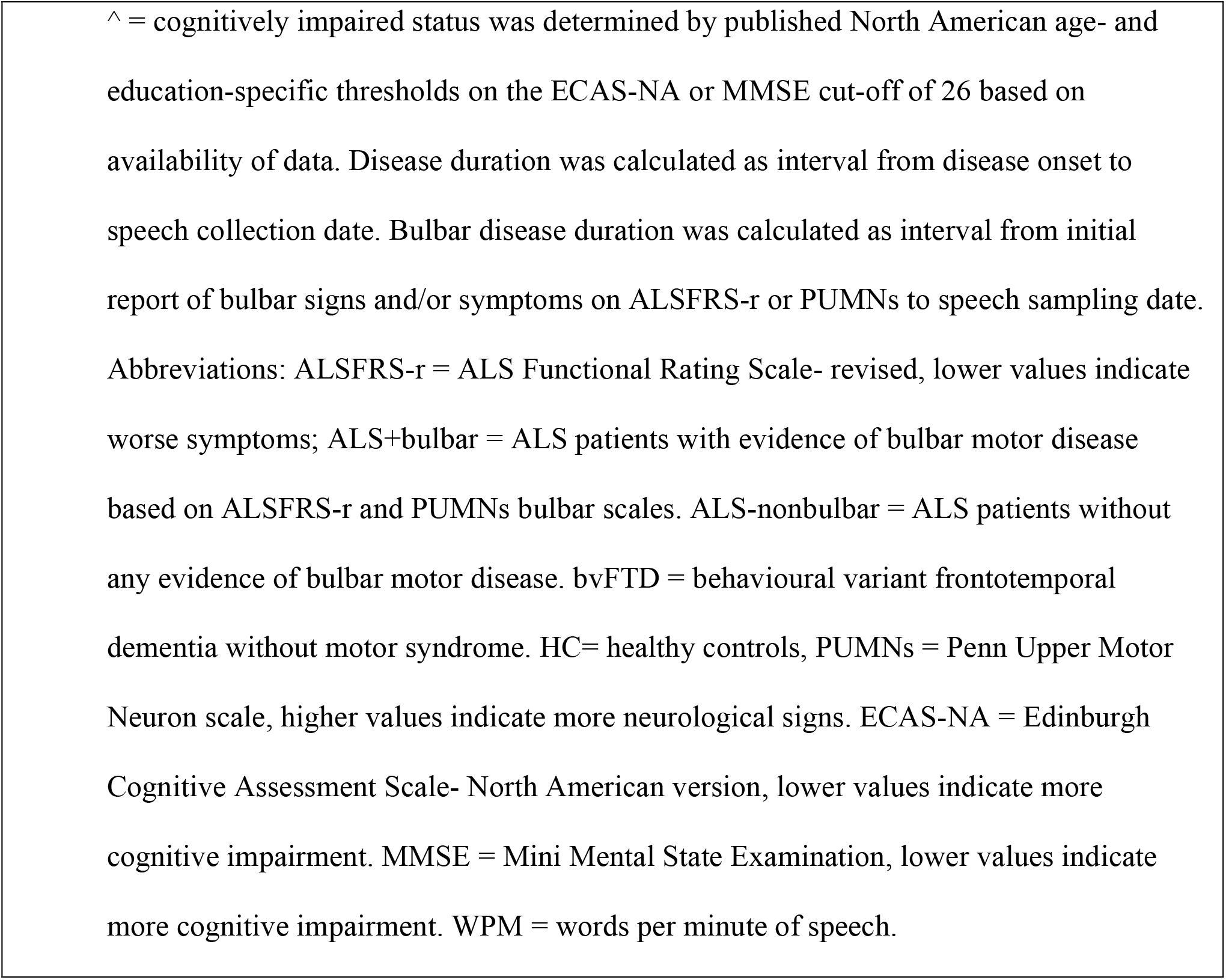
Demographics and clinical characteristics of participants. Normally distributed variables are summarized in mean (sd) and non-normal variables in median [IQR].

### 2.3. Digital speech data collection and processing

All participants provided a natural speech sample by describing the Cookie Theft picture scene from the Boston Diagnostic Aphasia Examination.^25^ A subset of the ALS patients (n = 24, 34%) provided a second picture description on the same day, elicited by the picnic scene from the Western Aphasia Battery.^26^ Participants were asked to describe the picture(s) in as much detail as they could. The speech samples were recorded on a single channel without compression at a sampling rate of 16 KHz and bit depth of 16 in a quiet room with minimal background noise. Groups did not differ in average sample duration (79±20 seconds).

Recordings were transcribed by trained transcribers and processed with automated algorithms to derive acoustic measures of tongue movement size and speed (described below).

We also calculated speaking rate (WPM) as total words divided by total time (including pauses), as it is a commonly used marker of bulbar motor disease in ALS.

### 2.4 Automatic vowel acoustic analysis

Acoustics pertain to the physical properties of sounds. Specifically, we analyze formant frequencies which are the natural resonant frequencies of the vocal tract based on the position of the tongue^27^, Vowel acoustics, specifically formant frequencies, are strongly related to tongue movement during articulation:^27^ the first formant, F1, is associated with the height of the tongue in the mouth during speech; the second formant, F2, is associated with the anterior and posterior dimension of tongue position. Vowels were automatically processed and analyzed using the Forced Alignment & Vowel Extraction (FAVE) system.^28^ First, FAVE aligned the acoustic signal to the text transcript, creating a Praat TextGrid file^29^ labeled at the phoneme (i.e., consonant and vowel) and word levels. FAVE’s automatic phoneme-level segmentation shows high congruency with gold-standard manually defined boundaries.^30^ Next, FAVE tagged all vowels with the CMU word pronunciation dictionary^31^ and extracted duration and formant values for each vowel token. F1 and F2 were extracted at the “steady-state” of each vowel token (approximately mid-point of the vowel when it is most stable), and at periodic increments through the vowel trajectory (at 20%, 35%, 50%, 65%, and 80% of vowel duration). We only analyzed stressed vowels, which were assigned by the pronunciation dictionary, and analyzed all available words except fillers, i.e., “um” and “uh”. bvFTD group produced fewer vowels on average than ALS groups (Table 1) which we adjusted for in the models.

Formant values in the logarithmic Hz scale were transformed to the linear Bark scale using [(26.81*F)/(1960+F)] - 0.53, where F= frequency in Hz.^32^ The Bark scale is perceptually uniform and is defined so that the critical bands of human hearing each have a width of one Bark. Bark scaling is beneficial in studies of dysarthria as it more closely corresponds to the perceptual organization of speech sounds in the auditory system, facilitating the interpretation of results in terms of speech perception and intelligibility. Bark transformation has shown to reduce the absolute differences between healthy females’ and males’ VSAs and preserve strong relationships with intelligibility in studies of dysarthria.^33^

#### 2.4.1. Articulatory-acoustic vowel measures of size and speed

Articulatory-acoustic measures that previously demonstrated sensitivity to bulbar dysfunction in ALS from structured speech tasks were adapted for natural speech^34,35^. When both picture descriptions were available for a participant, vowels were pooled across the two pictures as the average vowel durations and formant values did not differ between the two picture stimuli (*p*>0.96, repeated-samples t-tests). We calculated two articulatory-acoustic measures per participant:

1. Vowel space area (VSA): the 2-dimensional area of the triangle in F1xF2 space, in Bark^2^, formed by the three “corner” vowels, which are vowels produced at the extremes of tongue displacements. VSA serves as an acoustic proxy for a speaker’s articulatory working space, or tongue movement area.^35^ The area is calculated between the following vowels at steady-state: /i/ as in “cookie”, /α/ as in “hot”, /u/ as in “stool,” and /ʊ/ as in “cookie”, using F1_/i/_ * (F2_/α/_−F2_/u+ʊ/_) + F1_/α/_ * (F2_/u+ʊ/_−F2_/i/_) + F1_/u+ʊ/_ * (F2_/i/_−F2_/α/_)/2. As denoted, we combined /u/ and /ʊ/ to form a single vowel class representing the highest, back-most vowel, as these two vowels largely overlapped in acoustic space, and were both produced half as many times as the other two vowels, /i/ and /a/. A median of 6 vowel tokens were produced per vowel class per recording. We calculated the triangular VSA created between the centroids of these three vowel classes, i.e., mean F1 and F2 across vowel tokens within a vowel class (see graphical display of VSA in Supplementary Materials A).
2. Mean F2 slope: the rate of change of F2, in Bark/ms, averaged across all vowel trajectories, i.e., every stressed consonant-vowel-consonant transition. Slopes were calculated for each vowel token using F2_max_ -F2_min_/vowel duration, across 20-80% of the vowel trajectory, then averaged per person. F2 slope is an acoustic measure of the rate of change by the tongue in the front-back axis, the primary articulatory gesture associated with consonant-vowel-consonant transitions.^36^ A shallower slope indicates longer durations and less movement excursion.

### 2.5. Listening task and perceived listener effort

A random subset of Cookie Theft recordings from patients with ALS (n = 40, 56% of the total group) were rated perceptually for “listener effort”, a construct of speech intelligibility that is often described as the perceived cognitive resource necessary for speech recognition.^37^ The subset with listener effort data did not differ from the whole ALS group in demographics or ALSFRS-r bulbar score. Listener effort is more sensitive to early speech changes than intelligibility, which remains intact at the expense of increased effort of listeners.^38^

Listener effort was rated via REDCap, an online survey platform. Two independent listeners (S.S. and S.A.) who were blinded to the bulbar group designation were asked about their perceived effort of listening to 30-second clips of the Cookie Theft recordings (mean duration = 29.07 ± 0.16 seconds). S.S. selected the audio clips manually by arbitrarily selecting the largest section with the most voicing as seen on the waveform, which is blinded to articulatory information. Listeners were asked to answer the following question: “How effortful was it for you to understand the speaker? Remember, we are asking how hard you worked, not how well you did.” Answers were recorded on a visual analog scale, which consisted of a slider on an unmarked scale that the rater could position freely, with end points labeled “very” (converted to a score of 100) and “not at all” (score of 0).^39^

Inter-rater agreement for listener effort was good (ICC = 0.72, *p*<0.001) between the two raters, established by the intraclass correlation coefficient (ICC) on a mean-rating, consistency, 2-way random-effects model. A subset of recordings which was rated twice by each rater (24 samples, 20%). The intra-rater reliability (i.e., consistency within each rater) showed moderate to good consistency (ICC = 0.84 (S.S.) and 0.72 (S.A.), both *p*<0.001) on a single-rating agreement 2-way mixed-effects model. The average listener effort rating between the two raters was used for analysis.

### 2.6. Neuroimaging methods

High resolution T1 volumetric brain MRI scans were available for 18 ALS patients (9 ALS+bulbar, 11 ALS-nonbulbar) collected on a Siemens 3T Trio scanner within 1 year of speech sampling (10 ± 6.56 months). Due to the small sample size, we tested associations across both ALS groups. We statistically adjusted for the ALS group and time between MRI and speech in our models. The MRI subset did not differ from the whole ALS group in demographics or disease severity (ALSFRS-r total scores). We also confirmed that the bulbar group stratification at speech sampling matched the condition at MRI, i.e., none of the ALS-nonbulbar patients had developed bulbar signs or symptoms at the time of MRI scan, and all ALS+bulbar patients had evidence of bulbar disease at the time of MRI. The clinical motor scores remained the same for all but two patients, for whom there was a 2-point progression on the ALSFRS-r bulbar scale from speech sampling to their next closest MRI dates.

Briefly, T1-weighted images were preprocessed using antsCorticalThickness.sh (https://github.com/ftdc-picsl/antsct-aging^40^). The Schaefer 400 parcellation^41^ was warped to the individual T1, which divides each hemisphere into 200 roughly equal-sized areas. Mean cortical thickness values were extracted from each label. Average cortical thickness for the bilateral orobuccal homunculus (oral primary motor cortex (PMC)) and hand PMC regions were calculated by combining values across anatomically pre-defined Schaeffer labels relating to tongue motion and finger tapping motion in healthy controls on task-based fMRI^42^. The oral PMC region was composed of label numbers 43, 45, and 47 (left hemisphere), and labels 245, 246, and 248 (right hemisphere). The hand PMC region was the average of labels 53, 57, and 60 (left hemisphere) and labels 255, 256, and 259 (right hemisphere).^42^

### 2.7. Statistical Considerations

Vowel measures were compared between ALS+bulbar, ALS-nonbulbar, bvFTD, and HC groups using analyses of covariance (ANCOVAs) adjusting for group differences in age and total vowels (Table 1). We conducted pairwise comparison of means using the Tukey method for the Group factor, which adjusts for pairwise comparisons and takes into account the effects of the covariables. We tested group differences between ALS+bulbar and ALS-nonbulbar after adjusting for %FVC and ECAS ALS-Specific scores.

We examined associations with bulbar motor severity in ALS+bulbar patients using a sum of ALSFRS-r bulbar and PUMNs bulbar scores (summed bulbar score). Lower values on ALSFRS-r but higher values on PUMNs indicate more bulbar disease; therefore, we reversed the PUMNs so a maximum summed bulbar score of 16 indicated absence of all bulbar signs and symptoms. Four ALS+bulbar patients without PUMNs scores were omitted from this analysis. Linear regression models adjusted for ECAS ALS-Specific score and %FVC. As these not significant covariables and were only available in a subset of patients, we report Pearson’s correlations in the manuscript and full model results in Supplementary Materials B.

We also examined relations with perceived listener effort, serving as a parameter of communication limitation largely related to the speaker’s intelligibility.^37^ We compared 17 ALS speakers requiring zero listener effort vs. 23 speakers rated >0 using t-tests, and tested associations with listener effort in the 23 ALS speakers rated >0 using linear regressions, adjusting for ALS-Specific score and %FVC. We report Pearson’s correlations here, and full model results in Supplementary Materials B.

To test our hypothesized independence from cognitive and respiratory deficits, we examined Pearson’s correlations with ECAS ALS-Specific scores and %FVC in the ALS patients. We conducted identical analyses as above with speaking rate as a routinely-used comparative measure of bulbar motor disease. Supplementary Materials B reports all tested clinical correlations.

We examined associations with cortical atrophy in oral PMC and hand PMC in ALS using linear regressions, adjusting for ALS subgroup and time between MRI and speech recording. All statistical tests were two-tailed with a significance threshold of α = 0.05. Statistical analyses were performed with R version 4.1.0 and RStudio version 1.4.1717.

### 2.8. Standard Protocol Approvals, Registrations, and Patient Consents

The Institutional Review Board of the Hospital of the University of Pennsylvania approved the study of human subjects, and all participants agreed to participate in the study by informed written consent.

## 3. RESULTS

### 3.1. VSA and F2 slope are exclusively reduced in ALS+bulbar

Figure 1 shows the vowel measures for the four groups. Patients with ALS+bulbar had significantly smaller VSA than ALS-nonbulbar (Tukey’s adjusted *p*=0.0089, |*d*|=0.94), bvFTD (*p*<0.001, |*d*|=0.89), and HC (*p*=0.014, |*d*|=0.73). ALS+bulbar had significantly shallower F2 slopes than ALS-nonbulbar (*p*=0.0026, |*d*|=1.04), bvFTD (*p*<0.001, |*d*|=1.47), and HC (*p*=0.0015, |*d*|=0.99). The two vowel measures were not reduced in ALS-nonbulbar nor in bvFTD compared to HC, *p* > 0.53. Age and total vowels were not significant covariables for either measure ALS+bulbar vs. ALS-nonbulbar group differences remained significant after adjusting for %FVC and ALS-Specific scores, and neither were significant covariables (see Supplementary Materials B for post-hoc linear regression model results).

**Figure 1.**
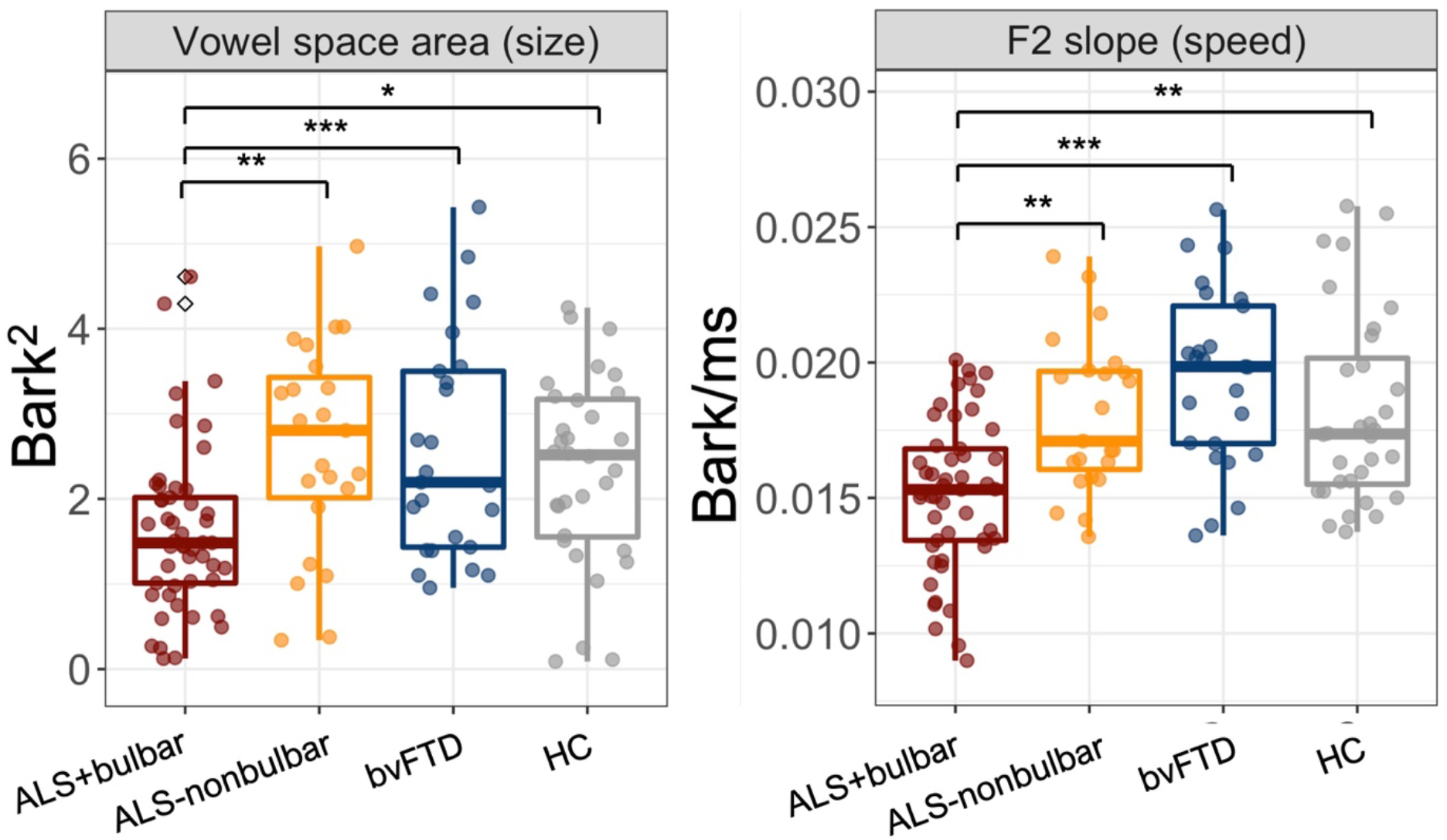
Vowel space area (measure of tongue movement size) and mean F2 slope (measure of speed) are significantly reduced in ALS patients with bulbar motor disease (ALS+bulbar) compared to ALS patients without bulbar disease (ALS-nonbulbar), behavioural variant frontotemporal dementia (bvFTD), and healthy controls (HC). All other contrasts were non-significant, p>0.05. *=p<0.05, **=p<0.01, ***=p<0.001; Tukey’s adjusted HSD.

By contrast, speaking rate was reduced in both ALS+bulbar and bvFTD groups, compared to ALS-nonbulbar and HC (Table 1).

### 3.3. VSA and F2 slope correlate with bulbar scores but not cognitive or respiratory scores

Figure 2 shows the significant relationship between vowel measures and summed bulbar scores in ALS+bulbar. A lower summed bulbar score, which indicates greater bulbar motor disease, was significantly correlated with a smaller VSA, R=0.33, *p*=0.043, and shallower F2 slopes, R=0.38, *p*=0.011. ECAS ALS-Specific scores were not significant covariables in either model (Supplementary Materials B).

**Figure 2.**
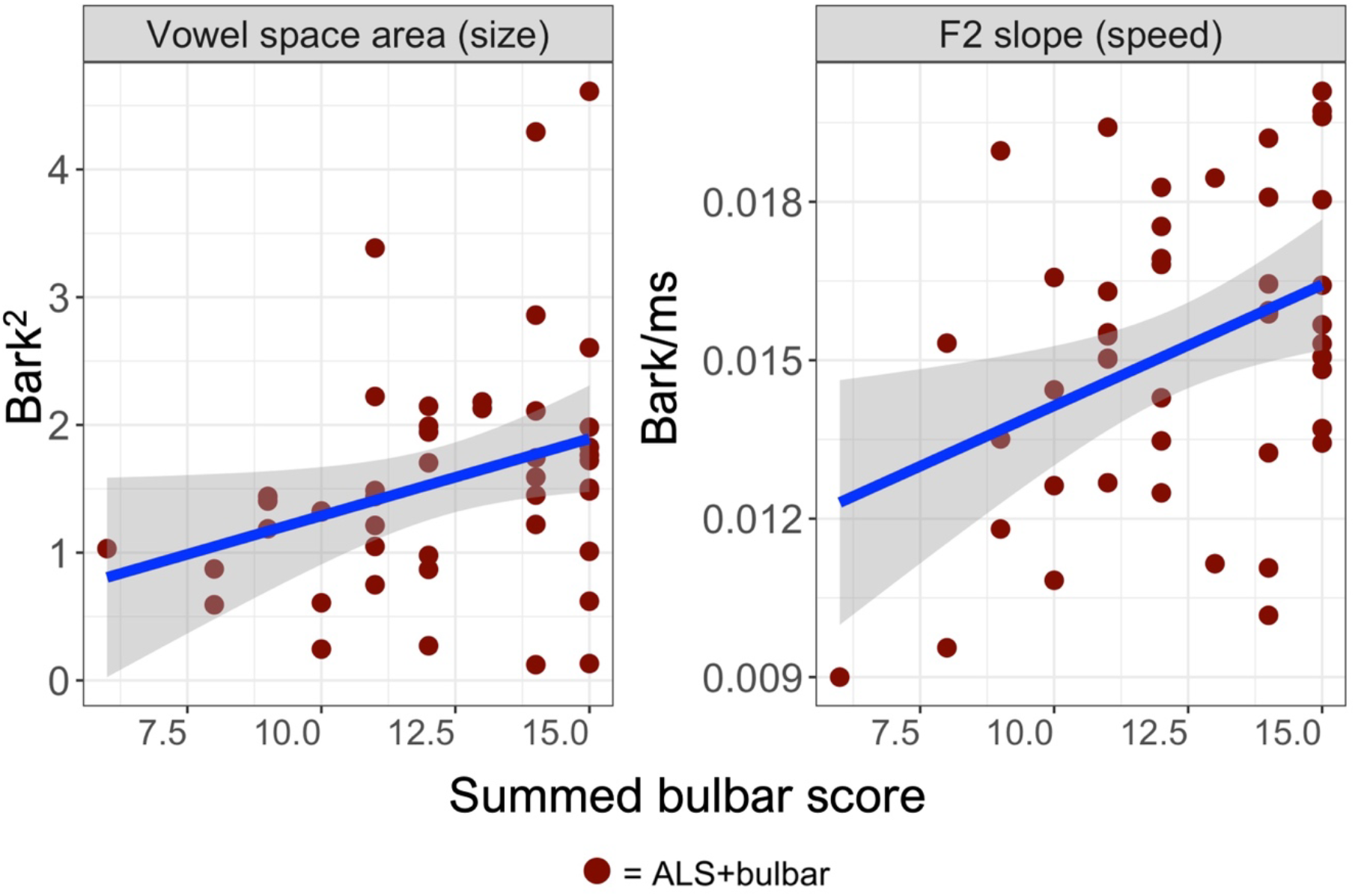
Worse bulbar disease (lower summed bulbar scores composed of ALSFRS-r bulbar and PUMNs bulbar scores) is significantly associated with smaller vowel space area and shallower F2 slopes in ALS+bulbar patients.

Neither vowel measure correlated with %FVC (*p*>0.20) nor ALS-Specific scores (*p*>0.80). Speaking rate did not correlate with summed bulbar scores (*p*=0.10) but rather was significantly associated with %FVC, R=0.31, *p*=0.009, and ECAS ALS-Specific scores, R= 0.42, *p*=0.0007 (Supplementary Materials C).

### 3.4. VSA correlates with listener effort

VSA was significantly reduced in ALS patients with perceived listener effort >0 compared to patients requiring 0 listener effort (*p*=0.041, |*d*|=0.45, plot not shown). Greater perceived listener effort was associated with smaller VSA, R= -0.43, *p*=0.041 (Figure 3). ECAS ALS-Specific score was not a significant covariable in the model (Supplementary Materials B).

**Figure 3.**
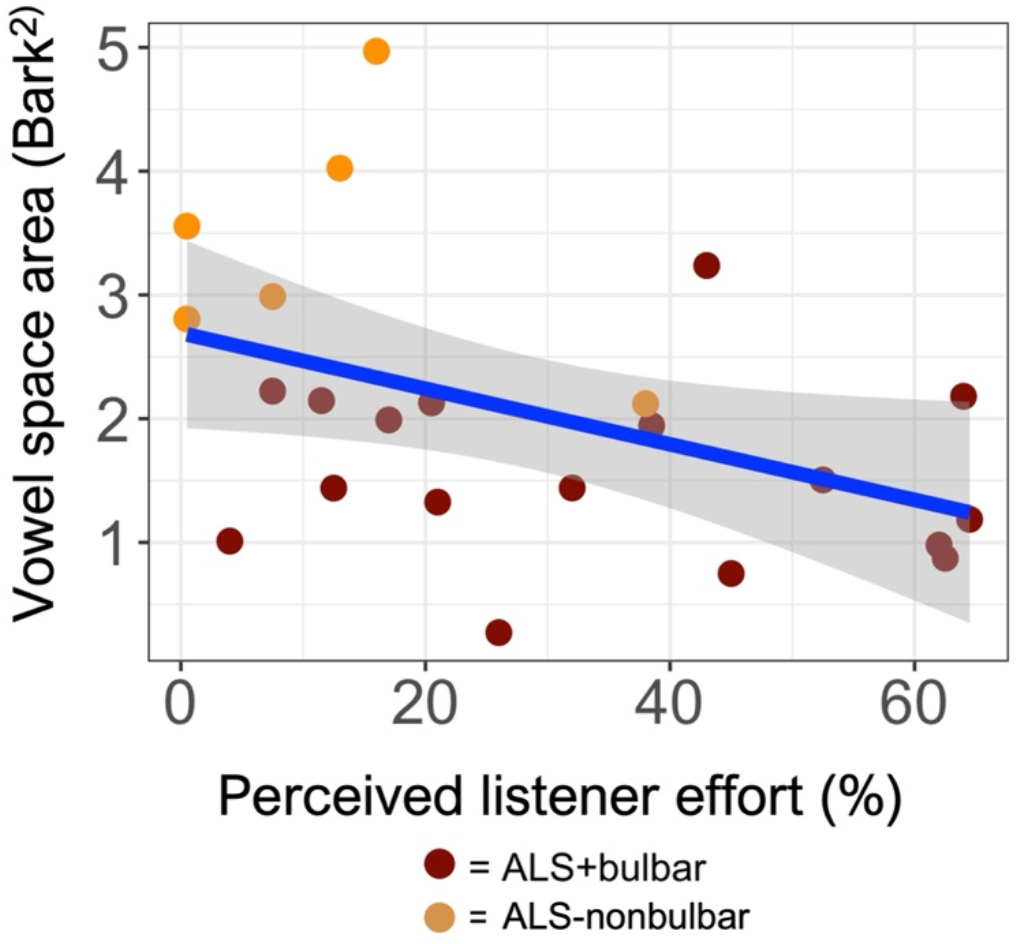
Greater perceived listener effort is significantly associated with smaller vowel space area in ALS.

F2 slope did not differ between listener effort groups nor relate to listener effort, *p*>0.17. Speaking rate was not associated with listener effort, *p*=0.30 (Supplementary Materials C).

### 3.5. F2 slope correlates with cortical thickness of the orobuccal homunculus of the primary motor cortex but not hand motor area

Reduced F2 slope was significantly related to cortical thinning in oral PMC in ALS after adjusting for time interval between MRI and speech sampling and ALS group, but not with hand PMC (Figure 4). Full model results are in Supplementary Materials B. VSA was not associated with cortical thickness in either motor region, *p*>0.62.

**Figure 4.**
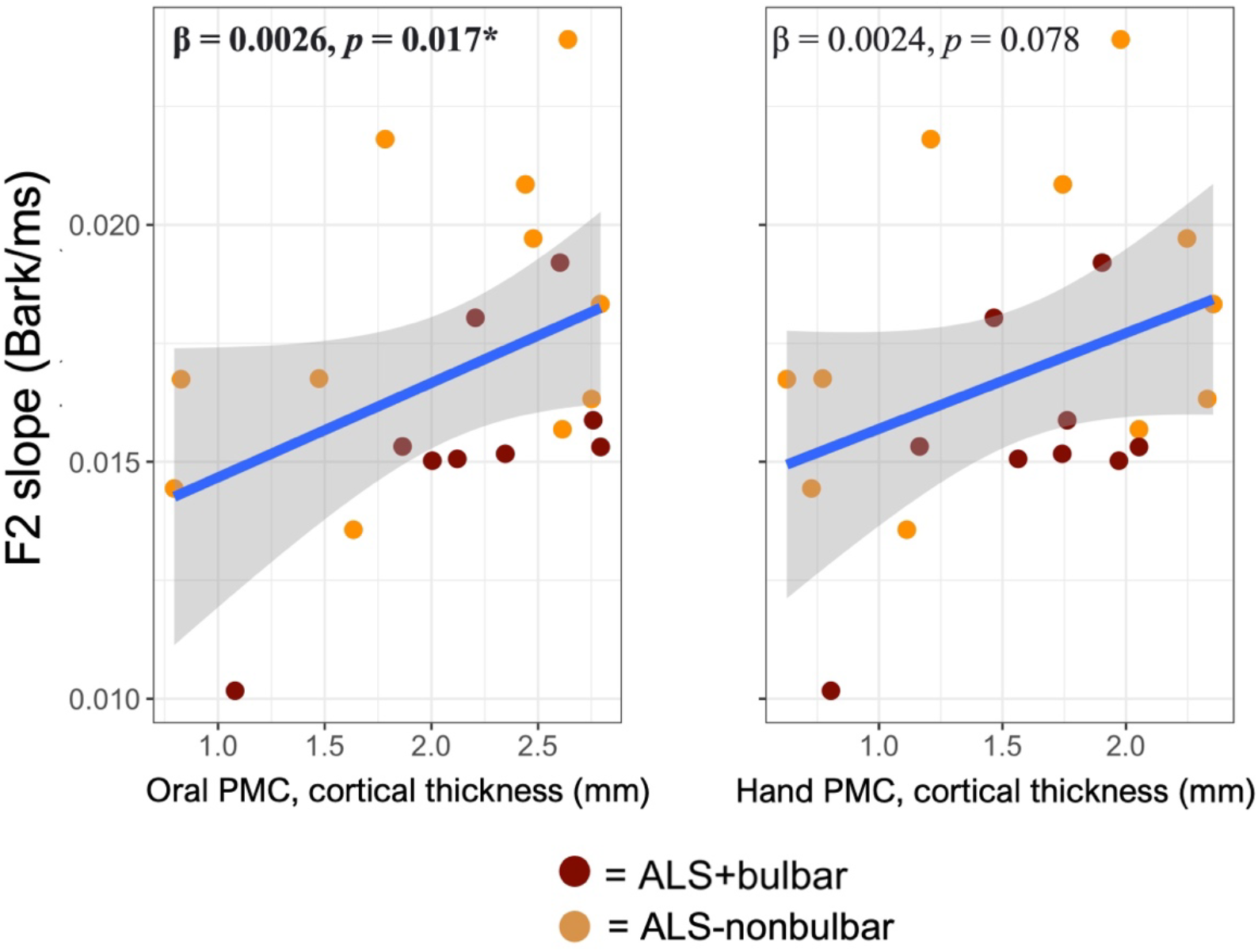
Shallower mean F2 slope was significantly related to greater cortical thinning in orobuccal homunculus, i.e., oral primary motor cortex (oralPMC) (left panel). Oral PMC consisted of average thickness across 3 left and 3 right hemispheric regions previously found to correspond with tongue motion in healthy controls. F2 slope was not significantly related to atrophy in hand PMC (right panel). Beta coefficients and p values for each ROI are noted.

## 4. DISCUSSION

Given the devastating impact of bulbar motor disease on vital speech and swallowing functions, developing objective clinical markers that can reliably support diagnosis and tracking of bulbar motor disease is crucial in patients with ALS-FTSD, who present with mixed motor, respiratory, and cognitive impairments. In this study, we provide clinical validation data for vowel measures of natural speech that reflect tongue movement size (vowel space area) and speed (F2 slope), which are sensitive to bulbar motor disease in ALS based on multiple findings. Firstly, both measures were impaired only in ALS patients with evidence of bulbar motor signs or symptoms, compared to ALS patients without bulbar motor impairment, bvFTD, and healthy controls. Secondly, vowel measures declined with worse bulbar motor disease. Lastly, vowel space area was related to perceived listener effort and F2 slope was related to atrophy in orobuccal homunculus areas. Importantly, neither vowel measure was related to cognitive or respiratory impairment.

Our study found that vowel-based measures are sensitive indicators of neurodegenerative bulbar changes in ALS. Our findings are consistent with previous studies of structured speech tasks reporting smaller VSA and shallower F2 slopes in ALS patients with bulbar disease, which was related to reduced speech intelligibility and slower and smaller tongue movements.^43–45^ But traditional methods of measuring vowels are time-consuming and require expert training limiting their use as clinical markers, as vowel detection and formant extraction require hand-labelling the speech signal. Fortunately, recent advances in automatic algorithms such as FAVE employed in this study, have made it possible to analyze large samples of natural speech more efficiently. Natural speech may be a better measure of articulatory impairment than structured speech tasks, which are commonly used in existing digital speech assessment tools but may be less sensitive to subtle neurodegenerative changes. A study comparing natural speech (monologues) to structured speech tasks (sentence repetitions and passage readings) in Parkinson’s disease found that the former captured more articulatory deficits, likely due to its high face validity with real-life communication.^46^ Our findings and previous studies highlight the sensitivity of vowel-based measures to ALS motor disease processes ^47^, and future comparisons of natural vs. structured speech in ALS patients could further inform the task specificity of motor speech control and validate our approach. Our results represent a significant step toward the development of a fully automated tool for the large-scale evaluation of articulatory impairment during natural speech due to neurodegeneration.

Our results highlight the clinical specificity of vowel measures in detecting motor impairment in ALS, in comparison to speaking rate which is commonly used to assess bulbar motor disease. We found that vowel measures were selectively impaired in ALS patients with bulbar motor disease, whereas speaking rate (WPM) was also affected in non-motor patients with cognitive impairment (bvFTD). Correlational analyses indicated that vowel measures declined due to bulbar motor disease and resulted in greater listener effort, but were not related to respiratory or cognitive impairment. In contrast, speaking rate declined due to multiple motor and cognitive sources of impairment in ALS, and was not related to listener effort. Thus, speaking rate may not be a suitable motor-specific marker in patients with concomitant cognitive impairments, while vowel measures may offer greater diagnostic and tracking precision as a clinical tool. Future investigations should test the responsiveness of vowel measures in early or presymptomatic stages of bulbar motor disease.

We found that smaller VSA was related to greater perceived listener effort, suggesting that a smaller tongue working space results in vowels being produced more centrally that are harder to distinguish from each other. On the other hand, tongue movement speed, as measured by mean F2 slope, did not show a similar relationship to listener effort. This can be explained by previous studies in ALS that found a reduction in articulation rate prior to a decline in speech intelligibility, suggesting that slow speech may be a compensatory strategy used by patients to maintain intelligibility.^48^ VSA may also be particularly sensitive to swallowing impairment, specifically in the oral stage where tongue movements play a crucial role in bolus formation and manipulation. Studies in stroke patients have shown a strong link between VSA and swallowing efficiency.^49^ Given the importance of tongue working space to both speech and swallowing, further research should examine the relationship between VSA and swallowing parameters in ALS.

Our imaging analyses revealed a moderately strong relationship between tongue articulation speed (F2 slope) and cortical thinning in the average bilateral oral primary motor areas that innervate tongue muscles. However, we did not find a similar relationship with tongue movement size (VSA). Slow articulation is a cardinal feature of spastic dysarthria caused by upper motor neuron loss,^50^ while reduced tongue movement size caused by muscle weakness is a feature of flaccid dysarthria related to lower motor neuron loss in the brainstem. This may partially explain the lack of association with VSA in our exploratory MRI analysis, which was limited to the motor cortex. Since the availability of MRI data was limited, we analyzed both ALS+bulbar and ALS-nonbulbar patients together, despite their differing clinical presentations. Surprisingly, these two patient groups largely overlapped in the range of cortical thinning in oralPMC. However, some ALS-nonbulbar cases may have been misclassified due to missed subtle bulbar UMN signs, such as a brisk jaw jerk, which could explain why they have a relatively thin oralPMC and shallow F2 slopes. Indeed, chart review revealed that the four ALS-nonbulbar cases with the thinnest oralPMC developed bulbar UMN signs within 6 months from MRI, while the remaining ALS-nonbulbar cases did not develop bulbar disease for 9-36 months after MRI. Conversely, the ALS+bulbar cases may have more brainstem lower motor neuron involvement and a spared motor cortex, as lower motor neuron involvement in bulbar muscles has an overt presentation of atrophy, fasciculations, and “slurred” speech, which is likelier to be classified as ALS+bulbar. Future studies should aim to examine associations between articulatory-acoustic metrics and brainstem volumetrics to better understand the underlying neural mechanisms of speech production in ALS patients.

Analyzing natural speech tasks can be challenging due to the uncontrolled nature of the speech produced, which can result in diverse and unpredictable lexical content and varying amounts of speech data compared to structured speech. By using picture description as the task, we mitigate some of this variability and elicit topic-controlled speech with specific words that are produced by all speakers (e.g., “cookie”, “stool”, and “water”). Considering coarticulation effects of surrounding consonants on vowel formants, we maximized vowel data by pooling data from multiple picture descriptions when available. While these strategies were effective, additional speech tasks that elicit longer samples of natural speech, such as descriptions of a story elicited by a wordless picture book or conversations on controlled topics, could be valuable. These naturalistic tasks have the advantage of being easy to administer and executed by patients with varying degrees of cognitive impairment. The present study is also limited by small sample sizes, primarily due to the low availability of MRI and speech data in ALS patients with severe bulbar impairment. These patients often experience respiratory dysfunction and have difficulty tolerating the supine position necessary for MRI. To improve upon these limitations, future studies and collaborative efforts should be encouraged to collect speech data from the widest possible range of patients. Although this study demonstrated strong clinical validity of vowel articulation measures to bulbar motor disease in ALS, the repeatability (test-retest reproducibility) of the novel metrics should be established in future.

With these caveats in mind, we conclude that automatic vowel analysis of natural speech can provide highly objective and motor-specific measures of bulbar disease in patients with ALS spectrum disorders. The proposed measures are robust to cognitive and respiratory dysfunction, are clinically interpretable, and can be derived from a brief natural speech sample.

## Data Availability

All data produced in the present study are available upon reasonable request to the authors

