## Supplementary Materials for "Digital markers of motor speech impairments in natural speech of patients with ALS-FTD spectrum disorders"

**Supplementary Materials A - Graphical representations of vowel space area**

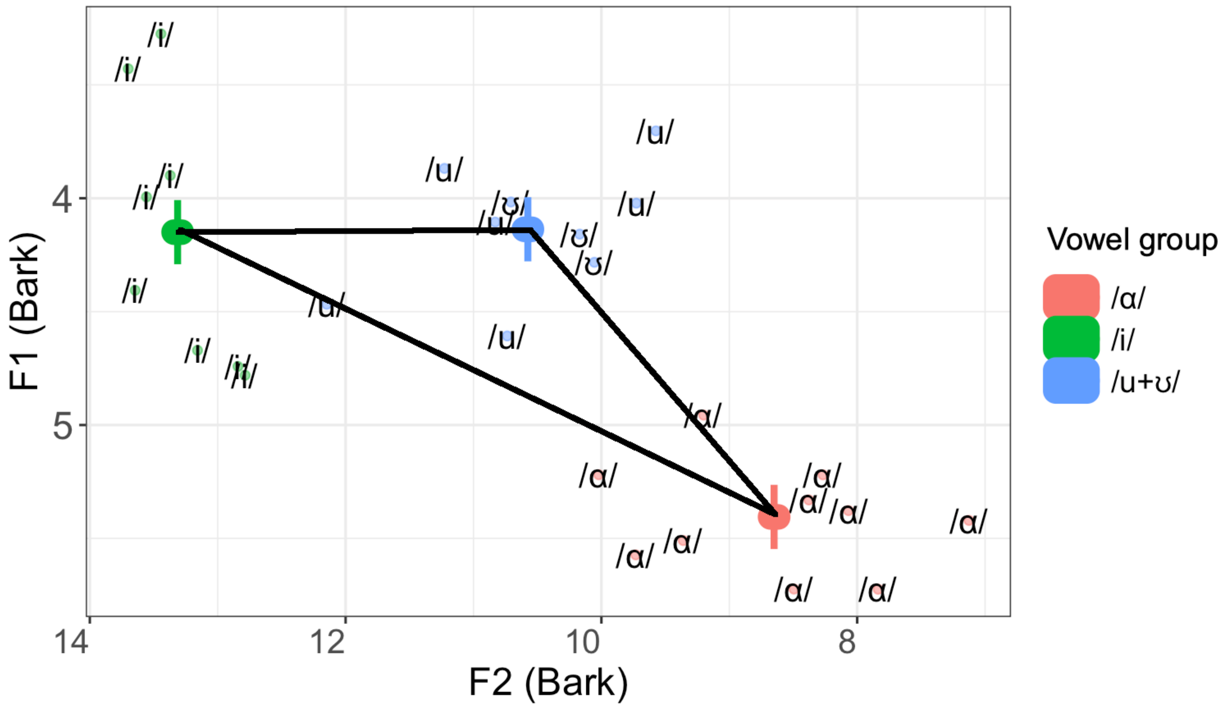


*Supplementary Figure 1.* Graphical representation of vowel space area in a single speaker, calculated as the triangular area between the three corner vowels in F1xF2 space. As shown, /u/ and /ʊ/ (in blue) largely overlap in acoustic space but are produced half as many times in the Cookie Theft as /i/ and /a/, thus were pooled together to represent a single high-back vowel class (in blue).

**Supplementary Materials B – Linear regression model results for clinical analyses**

*Supplementary Table 1.* Results of post-hoc linear regression models adjusting for potential respiratory and cognitive covariables on vowel measures in (A) ALS+bulbar vs. ALS-nonbulbar group comparisons; (B) correlations with clinical bulbar scores, and (C) correlations with perceived listener effort.

| | | | | |
|---|---|---|---|---|
| **(A) Group differences between ALS+bulbar and ALS-nonbulbar** | | | | |
| tVSA ~ Group + total vowels + age + %FVC + ALS-Specific score | | | | |
| | Estimate | Std.error | *t*-value | *p*-value |
| (Intercept) | 1.2762465 | 1.7416171 | 0.733 | 0.467 |
| Group (ref. = ALS+bulbar) | 0.8178743 | 0.3088343 | 2.648 | 0.0107* |
| Total vowels | 0.0062577 | 0.0032531 | 1.924 | 0.0599 |
| age | 0.0085972 | 0.015141 | 0.568 | 0.5726 |
| %FVC | -0.0006892 | 0.0067677 | -0.102 | 0.9193 |
| ALS-Specific score | -0.0092769 | 0.0157157 | -0.59 | 0.5575 |
| Mean F2 slope ~ Group + total vowels + age + %FVC + ALS-Specific score | | | | |
| (Intercept) | 0.01589808 | 0.0046233 | 3.439 | 0.001159 |
| Group (ref. = ALS+bulbar) | 0.0032316 | 0.00081983 | 3.942 | 0.000243*** |
| Total vowels | -3.901E-06 | 8.636E-06 | -0.452 | 0.65338 |
| age | 2.0688E-05 | 4.0193E-05 | 0.515 | 0.60894 |
| %FVC | -2.384E-05 | 1.7966E-05 | -1.327 | 0.190362 |
| ALS-Specific scores | 1.153E-06 | 4.1719E-05 | 0.028 | 0.978065 |
| **(B) Correlations with summed bulbar scores** | | | | |
| tVSA ~ Summed bulbar score + ALS-Specific score + %FVC | | | | |
| (Intercept) | -1.04327 | 1.77545 | -0.59 | 0.56 |
| Summed bulbar score | 0.1184 | 0.06974 | 1.7 | 0.047* |
| ALS-Specific score | 0.00931 | 0.01604 | 0.58 | 0.57 |
| %FVC | 0.00572 | 0.00784 | 0.73 | 0.47 |

| | | | | |
|---|---|---|---|---|
| Mean F2 slope ~ Summed bulbar score + ALS-Specific score + %FVC | | | | |
| (Intercept) | 0.01200814 | 0.00487913 | 2.46 | 0.02 |
| Summed bulbar score | 0.00044221 | 0.00019166 | 2.31 | 0.028* |
| ALS-Specific score | -8.85E-06 | 0.00004408 | -0.2 | 0.842 |
| %FVC | -1.975E-05 | 0.00002154 | -0.92 | 0.366 |
| (C) **Correlation with perceived listener effort** | | | | |
| tVSA ~ listener effort + ALS-Specific score | | | | |
| (Intercept) | -0.0831 | 1.8623 | -0.04 | 0.965 |
| Listener effort | -0.0178 | 0.01 | -1.78 | 0.044* |
| ALS-Specific score | 0.0302 | 0.0221 | 1.37 | 0.19 |

**Supplementary Materials C – Correlations with clinical scores for motor, respiratory and cognitive function**

*Supplementary Figure 3.* Scatterplots and Pearson's correlations for relationships between vowel measures (VSA and F2 slope) and speaking rate, and bulbar motor scores, perceived listener effort (%), respiratory capacity (%FVC), and cognitive scores (ECAS ALS-Specific scores). Bolded results indicate significant correlations.

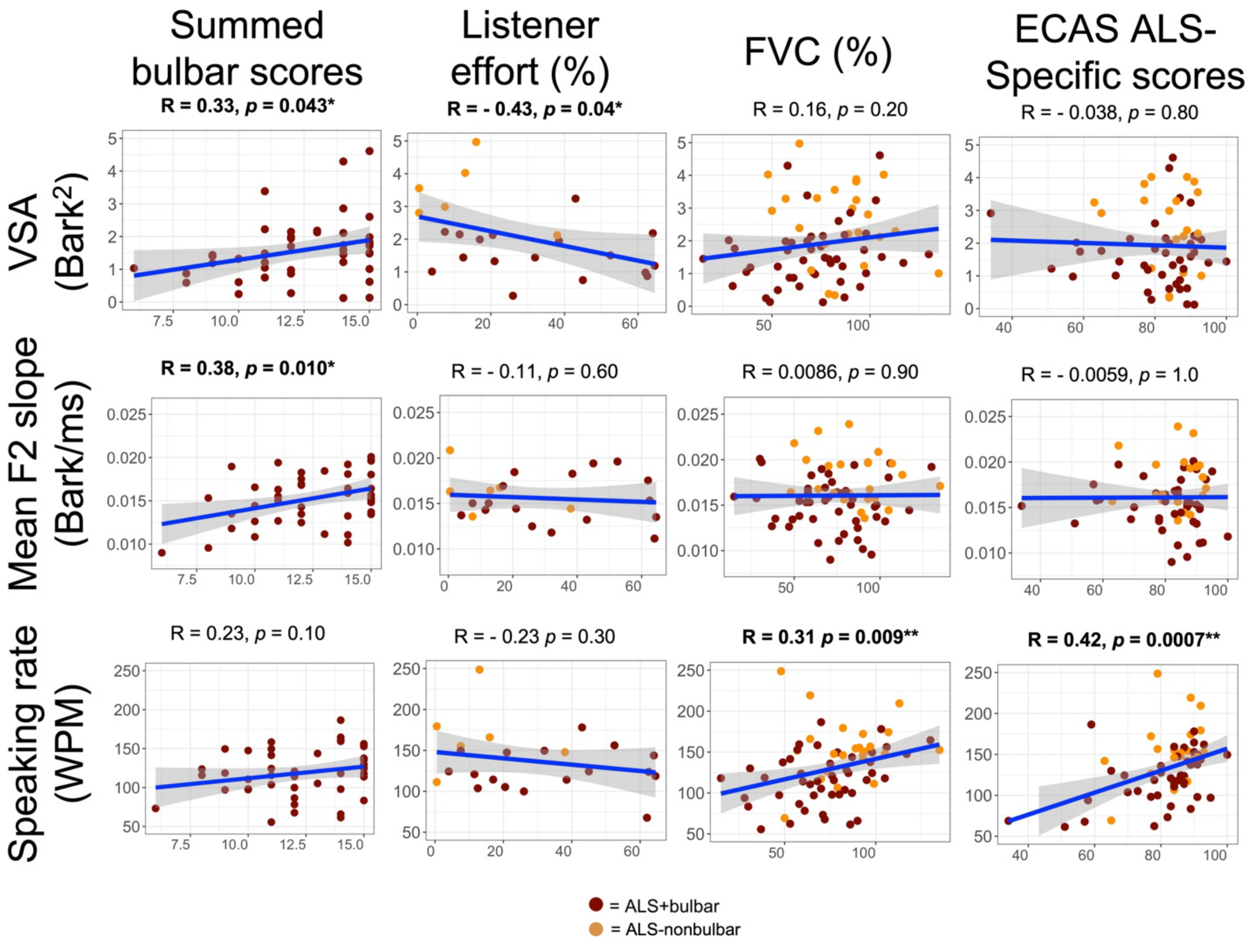

**Supplementary Materials D – Linear regression model results for neuroimaging analyses**

*Supplementary Table 2.* Linear regression model results testing associations between F2 slope and cortical thickness of oral and hand primary motor cortical (PMC) regions. Shallower F2 slope was associated with thinning in oral PMC, but not hand PMC.

| **F2 slope ~ Oral PMC cortical thickness** + time between MRI and speech + group | | | | |
|---|---|---|---|---|
| | Estimate | Std.error | *t*-value | *p*-value |
| (Intercept) | 0.0101069 | 0.0022155 | 4.56 | 0.00032 |
| Oral PMC cortical thickness (mm) | 0.0025777 | 0.0009724 | 2.65 | 0.01744* |
| Time MRI-to-speech (months) | -0.0000812 | 0.0001001 | -0.81 | 0.42951 |
| Group (ref = ALS+bulbar) | 0.0028443 | 0.00118 | 2.41 | 0.02833* |
| **F2 slope ~ Hand PMC cortical thickness** + time between MRI and speech + group | | | | |
| (Intercept) | 0.011902 | 0.002065 | 5.76 | 0.000029 |
| Hand PMC cortical thickness (mm) | 0.00238 | 0.001216 | 1.96 | 0.078 |
| Time MRI-to-speech (months) | -0.000067 | 0.000109 | -0.62 | 0.546 |
| Group (ref = ALS+bulbar) | 0.002521 | 0.001267 | 1.99 | 0.064 |
